# Mechanistic model calibration and the dynamics of the COVID-19 epidemic in the UK (the past, the present and the future)

**DOI:** 10.1101/2021.05.18.21257384

**Authors:** Harry Conn, Robyn Taylor, Mark J. Willis, Allen Wright, Victoria Bramfitt

## Abstract

▪ We augment the well-known susceptible – infected – recovered – deceased (*SIRD*) epidemiological model to include vaccination dynamics, implemented as a piecewise continuous simulation. We calibrate this model to reported case data in the UK at a national level,
▪ Our modelling approach decouples the inherent characteristics of the infection from the degree of human interaction (as defined by the effective reproduction number, *R*_*e*_). This allows us to detect and infer a change in the characteristic of the infection, for example the emergence of the Kent variant,
▪ We find that that the infection rate constant (*k*) increases by around 89% as a result of the B.1.1.7 (Kent) COVID-19 variant in England,
▪ Through retrospective analysis and modelling of early epidemic case data (between March 2020 and May 2020) we estimate that ∼1.2M COVID-19 infections were unreported in the early phase of the epidemic in the UK. We also obtain an estimate of the basic reproduction number as, *R*_0_ = 3.23,
▪ We use our model to assess the UK Government’s roadmap for easing the third national lockdown as a result of the current vaccination programme. To do this we use our estimated model parameters and a future forecast of the daily vaccination rates of the next few months,
▪ Our modelling predicts an increased number of daily cases as NPIs are lifted in May and June 2021,
▪ We quantify this increase in terms of the vaccine rollout rate and in particular the percentage vaccine uptake rate of eligible individuals, and show that a reduced take up of vaccination by eligible adults may lead to a significant increase in new infections.

## 1.0 Introduction

In the early stages of the COVID-19 pandemic (December 2019-March 2020) several papers were published reporting the development and use of mathematical models to forecast the potential infections and deaths which could arise unless stringent non-pharmaceutical interventions (NPIs) were introduced. These models proved to be vital in government decision making and so curtailing a major disaster (Adam, 2020).

In recent work (Willis, Wright, Bramfitt, & Diaz, 2021) we demonstrated that the well-known susceptible – infected - removed (*SIR*) and the susceptible – infected – recovered – deceased (*SIRD*) models^1^ (Bailey, 1975), are structurally equivalent to the model equations for a set of chemical reactions occurring in a well-mixed batch reactor in which the stoichiometry of the contagion reaction varies.

Due to this structural equivalence, we were able to exploit the sophisticated functionality of a commercial chemical engineering simulator that combines a dynamic modelling framework with kinetic regression tools. This allowed calibration of the characteristic rates of infection and removal of individuals and an estimate to be made regarding the effective reproduction number with respect to time. Our enhancement to the (*SIRD*) scheme was that we treat the time varying effective reproduction number *R*_*e*_ as a variable stoichiometric coefficient which responds exponentially to a change in NPI. We used temperature as a placeholder to develop this exponential variation. This choice was expedient as the modelling platform already had an Arrhenius equation (temperature dependent model) and regression tools for the equation coefficients. A significant advantage of this approach is that it decouples the inherent characteristics of the infection from the degree of human interaction *R*_*e*_ which is governed by the regime of NPIs in place at any time. This allows us to detect a variation in the disease transmission rate and infer a change in the characteristic of the infection, for example the emergence of the B.1.1.7 (Kent) variant.

Over the last twelve months several changes have occurred in the epidemics reporting, understanding, behaviour and treatment. Changes occurred and continue to occur for several reasons. Data reporting has changed as a result of increased testing, moving from targeted testing in hospitals to full-scale community testing, (BBC News, 2021). There have been changes in classification of data such as only reporting COVID-19 related deaths which occur within a 28 day time period of confirmed infection. In addition, the management and treatment of infected people has evolved resulting in an increase in recover rate and corresponding reduction in deaths. Of note, dexamethasone was found to reduce death rates in people hospitalized with COVID-19 on a ventilator and people receiving supplemental oxygen but not on a ventilator (The RECOVERY Collaborative Group, 2021).

Conversely, increase in infectivity and or morbidity has been reported for some emerging mutant viral strains, for example, the B.1.1.7 variant colloquially known as the Kent variant. More recently, vaccination roll out in a number of countries has been shown to reduce the probability an individual will become seriously ill following a COVID-19 infection (Scottish Government, 2020). Any model will have limited applicability unless it is possible to take account of these occurring changes.

In this paper we extend our modelling work (Willis, Wright, Bramfitt, & Diaz, 2021) examining the effects of COVID-19 in the UK. We apply our modelling approach to calibrate the characteristic rates of infection and removal of individuals and estimate the basic and effective reproduction numbers using historical UK data. Through piecewise model calibration we quantify the increase in transmission rate of the B1.1.7 (Kent) variant and obtain results comparable with those recently published (Briggs, 2021). We also use our calibrated model to provide a retrospective analysis of reported case data deficiencies in the early stages of the pandemic. Finally, we assess the UK Government’s roadmap for easing the third national lockdown as a result of the current vaccination programme (Elgot, 2021).

## 2.0 Methods

To calibrate our model, we use daily published UK Government (UK Government, Download the Data, 2021) and NHS England (National Health Service, 2021) data. This includes the numbers of new COVID-19 infections and deaths and the numbers of first and second vaccinations recorded each day.

### 2.1 Kinetic modelling

Defining *I* as an infected, *R* as a recovered and *D* as a deceased individual, the stoichiometric scheme describing the transition of individuals between the four compartments of an *SIRD* model is,

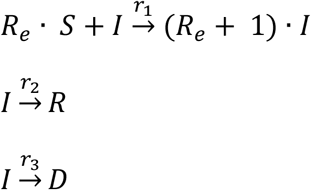

In this scheme, *R*_*e*_ is the (dimensionless) effective reproduction number. We observe that it is analogous to a stoichiometric coefficient in a chemical reaction scheme. The significant difference is that in chemical schemes, stoichiometric coefficients are constant whereas the effective reproduction number, *R*_*e*_ can vary throughout the course of an epidemic as NPIs are applied.

We extend this model to consider vaccination dynamics via,

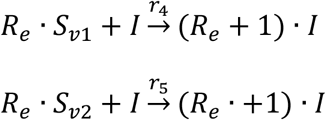

The two additional expressions account for the population subgroups of vaccinated individuals *S*_*v*1_ and *S*_*v*2_ becoming infected due to the limited immunity conferred by the vaccine. We develop a set of model equations by treating this scheme in the same way as a chemical scheme and applying the law of mass action kinetics (Willis, Wright, Bramfitt, & Diaz, 2021).

Defining, *n*_*S*_ [people] as the number of susceptible, *n*_*I*_ [people] the number of infected, *n*_*R*_ [people] the number of recovered, *n*_*D*_ [people] the number of deceased, *n*_*V*1_ [people] and *n*_*V*2_ [people] as the numbers of susceptible people that have received the first and second vaccine at rates *v*_1_ [people · d^−1^] and *v*_2_ [people · d^−1^] then the rate of change of the number of people in the various compartments of the model are,

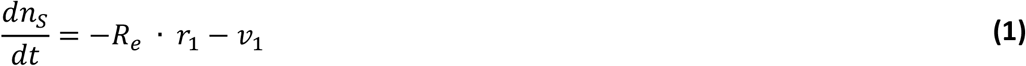

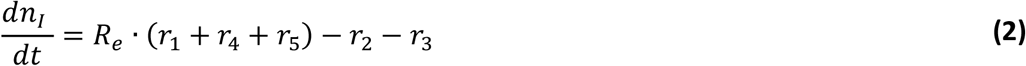

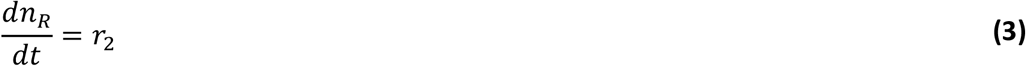

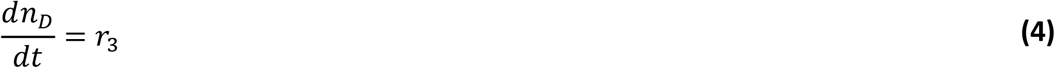

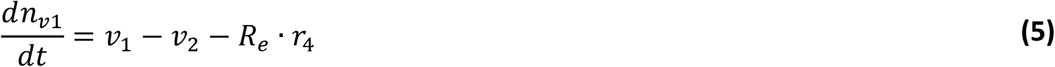

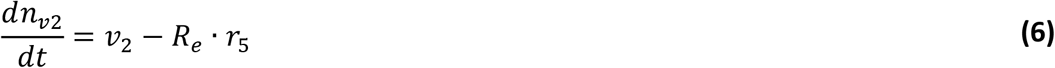

The additional rate terms [people · d^−1^] are,

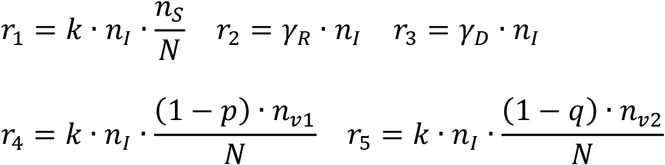

In these rate equations *k* [d^−1^] is the infection rate constant, *γ*_*R*_ [d^−1^] is the removal rate constant of recovered infectious individuals, *γ*_*D*_ [d^−1^] is the removal rate constant of deceased individuals, *N* [people] is the total population, *p* is the efficacy of the first vaccination and *q* the efficacy of the second. Substituting these terms into **(1)** to **(6)** gives,

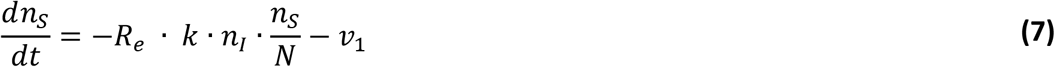

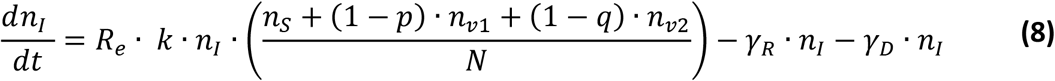

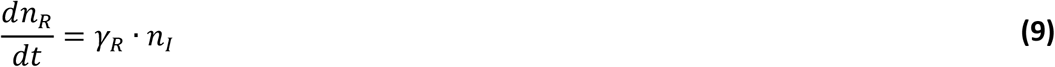

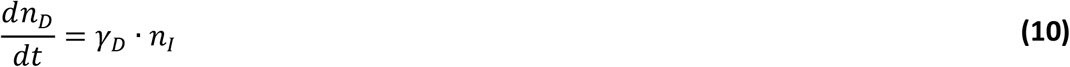

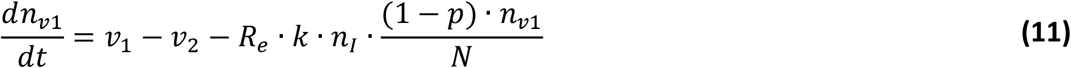

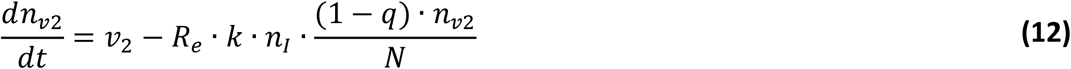

### 2.2 Modelling the variation in the effective reproduction number

In equations **(7)** – **(12)** we assume that *k* is constant and refer to this as the specific transmission probability per exposure time, a constant that is characteristic of the COVID-19 infection. It is assumed that *R*_*e*_ varies as an exponential function, and that the variation of *R*_*e*_ is due to measures taken as NPIs are changed during the epidemic. We use an Arrhenius equation to represent the exponential variation,

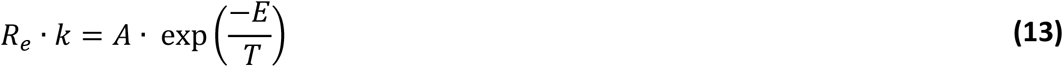

In this equation, *A* and *E* are constants which may be determined by model calibration at the same time as the other unknown parameters in equations **(7)** – **(12)**. In order to use this equation to capture variation in *R*_*e*_ we use temperature as a placeholder variable to represent the efficacy of NPIs. In other words, the model requires a sequence of temperature steps and ramps, which we use to represent the NPIs. As NPIs are changed the temperature can be adjusted to reflect the observed change in reported case data. The magnitude, direction and rate of the subsequent temperature changes in the model is representative of the stringency and efficacy of the NPIs. Increasing stringency of NPIs represented by decreasing temperature, relaxation of NPIs by increasing temperature.

### 2.3 Calculating the infection rate constant and the effective reproduction number

The number of infected individuals passes through a maximum (or minimum) at 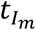, and at this point,

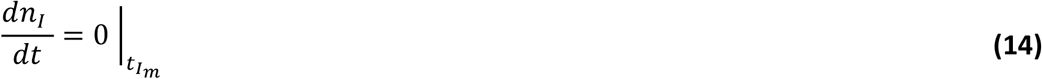

It follows that

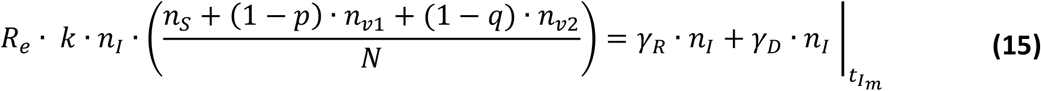

At this point is, *R*_*e*_ = 1, therefore the constant for specific transmission probability per exposure time can be calculated as,

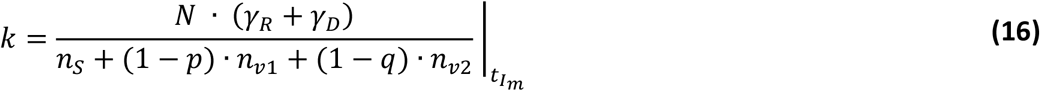

The value of the placeholder variable *T*(*t*) is known by inspection of the temperature profile imposed on the model. This allows the instantaneous effective reproduction number *R*_*e*_(*t*) to be calculated using **(13)**. The same analysis can be applied at any maximum or minimum (i.e. any point where,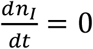) which is caused by NPIs being relaxed and tightened, therefore, we can use our model to detect any changes in the infection rate constant.

### 2.4 Control of disease transmission rates

The application of NPIs and the effective rollout of a vaccination programme can be used to control disease transmission rates and reduce the number of daily infections and deaths. NPIs reduce the effective reproduction number in order to exert control, whereas vaccination reduces the numbers of susceptible individuals.

Rearranging **(8)**, we have

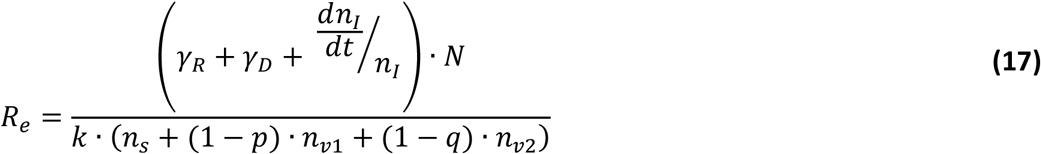

As any vaccination programme is rolled out, given that there will be less susceptible people, the value of *R*_*e*_ which will either maintain or decrease the cumulative number of infections (i.e. 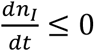) will increase above one and the maximum values is,

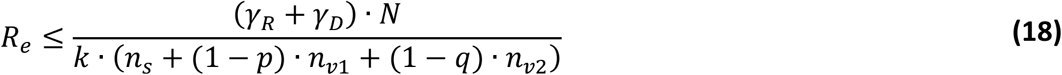

#### 2.4.1 Herd immunity thresholds

In equation (**18)** the fraction of susceptible people at any given time is,

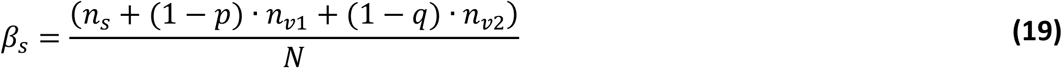

Defining, *β*_*R*_= 1 − *β*_*s*_, as the required fraction of the immune population, we can re-arrange **(18)** to give,

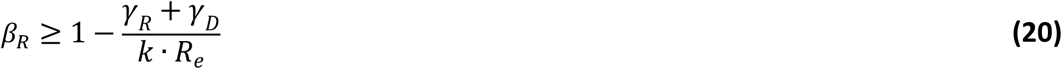

Typically, equation **(20)** is used in conjunction with the basic reproduction number at the onset of an epidemic to calculate a herd immunity threshold, ie the proportion of a population which must be removed from the susceptible population group in order for herd immunity to be achieved. In which case, if *γ*_*R*_ + *γ*_*D*_ ≈ *k* we have,

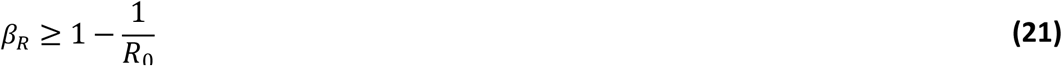

Many estimates of the value of *R*_0_ are available in the literature and are typically given in the range *R*_0_∼3.0 − 3.9 (Locatelli, Trächsel, & Rousson, 2021) (Lui, Tang, & Lam, 2020) (Bruce, et al., 2020). In other words, the required fraction of the population to achieve herd immunity is, *β*_*R*_∼0.67 − 0.75.

#### 2.4.2 Assessing compliance with the herd immunity threshold

Given the current vaccination roll-out in the UK, a conservative estimate of the fraction of the population that are immune at any time is,

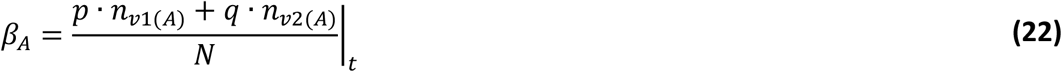

In this equation *n*_*v*1(*A*)_and *n*_*v*2(*A*)_ are actual cumulative numbers of individuals that have received a first and second vaccination. This estimate is conservative since we omit the population fraction who have recovered from infection. These people may have some immunity but are also eligible to be vaccinated.

Equation **(20)** is applicable at any time during the epidemic to calculate the required population immunity given an estimated value of *R*_*e*_. Therefore, as we move through the stages of the UK Government roadmap, equating equations **(20)** and **(22)** gives,

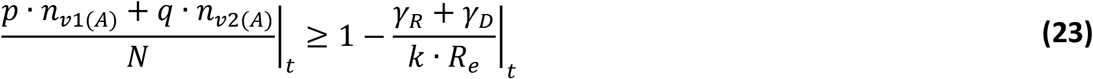

We can use equation **(23)** to estimate the current effectiveness of a vaccination programme using the UK Government’s roadmap for easing the third national lockdown (Elgot, 2021) in conjunction with calibrated model parameters (providing an estimate of the disease transmission rate) as well as an estimate of the effective reproduction number. A similar analysis has been recently presented by (Imperial College COVID-19 Response Team, 2021), (Mulheirn & Britto, 2021).

To estimate the current effectiveness of a vaccination programme, data to 12 May 2021 is used to calibrate the model and estimate *R*_*e*_. After this date, the UK Government’s roadmap for easing the third national lockdown (Elgot, 2021) is used for forecasting the future *R*_*e*_ profile where our estimated value of *R*_0_ is then used to represent the removal of all NPIs. To project ahead we use a vaccine forecast which assumes a total daily vaccination capacity.

Furthermore, on a given day, we assume that the number of second vaccinations corresponds to the number of first vaccinations given 12 weeks previously, and assume that all second vaccinations offered are taken up. The number of first vaccine doses given each day is calculated as the difference between the number of second doses given and the daily capacity. Finally, it is assumed that the number of first vaccinations are given until the number of eligible individuals is reached.

### 2.5 Model calibration

We use equation **(24)** as a configurable objective function to quantify the discrepancy between model response and reported case data. We use piecewise continuous numerical solution of the ODEs, coded in Simulink and integrated using *ode15s* algorithm for efficient integration of stiff ODEs. The solution is saturation limited between zero and the total population, and zero crossing is enabled. A sequential quadratic programming optimisation algorithm, *fmincon*^*2*^, minimises **(24)** by adjusting unknown model parameters.

The model parameters which may be optimised are the rate constants (*A, γ*_*R*_ and *γ*_*D*_), the initial number of infected individuals and the temperatures representing the sequence of NPIs. We use a pre-defined set of time values for the temperature co-ordinates which are chosen to be every 10 days to minimise the effect that weekend reporting has on model calibration^3^. The set of parameters is selected using the scripting system described in section 2.5.3 below.

The objective function is a weighted aggregate of the sum of the squared error between the reported values, *y*_*j*_^*^, and model predictions, *y*_*j*_, for the terms selected,

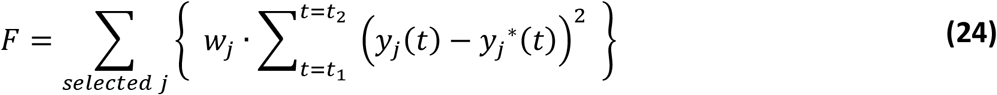

The model terms *y*_*j*_ that we use in the objective function are selected from cumulative deceased individuals, *n*_*D*_, the daily deaths, *r*_3_ and daily infection rate, *R*_*e*_ · *r*_1_. The summation is over the data window *t*_1_ to *t*_2_, and *w*_*j*_ is a weighting applied to a data set to account for differences in the magnitudes of individual variables. Data for deaths and daily deaths uses the actual date of death. Infected daily case data uses the date of reporting, and we apply a five day offset to account for the delay attributable to someone getting infected, presenting for testing, test analysis and reporting.

#### 2.5.1 Reported case data used for model calibration

In our earlier work (Willis, Diaz, Prado-Rubio, & von Stosch, 2020) (Willis, Wright, Bramfitt, & Diaz, 2021) and other previous work (Linka, Peirlinck, Sahli Costabal, & Kuhl, 2020) (Manenti, et al., 2020), the data selected for the objective function used cumulative figures for dead, infected and recovered individuals. We have observed that there were limitations in the accuracy of reported data, in particular for the number of recovered individuals to the extent that there was an apparent imbalance in the system. In this work we use the reported daily rates for infections and deaths and compare these with the equivalent terms calculated by the simulation. The use of the daily infection rate in the objective function is preferable for discrimination of calibrated model parameters because this particular model term is influenced by *k* only whereas the cumulative number of daily infections is influenced by *k, γ*_*R*_ and *γ*_*D*_. Furthermore, a characteristic of the epidemiological scheme is that there are only two pathways arising from infected individuals. Therefore, reported case data for one path (daily death rates) is sufficient for the other path (recovered individuals) to be inferred. In other words, it is not necessary to obtain data for numbers of recovered individuals to fully calibrate the kinetic parameters of the model.

#### 2.5.2 Data windows

As COVID-19 is a viral disease, it is prone to random mutations which have potential to change the characteristics of the disease, such as the rates infection, recovery and death (Public Health England, What do we know about the new COVID-19 variants?, 2021). These mutations have resulted in many stable variants of COVID-19 spreading throughout the global population. As numerous variants are present within the UK, the parameters of our calibrated model will represent an average of all those present. However, as shown by (Chowdhury, Scarr, MacAskill, & Marshall, 2021), the Kent variant of COVID-19 has been responsible for majority of new infections in the UK from November 2020 onwards.

Consequently, we calibrate our model over three distinct time periods.

Period one considers reported case data between the 12 June 2020 and 13 November 2020. We use this data to characterise the ‘original’ variant of COVID-19.

Period two is defined between 14 November 2020 and 24 March 2020 where the B.1.1.7 (Kent) variant is believed to dominate UK infections.

In period 3, we take a retrospective look at reported case data in the early stage of the epidemic considering data between March 2020 and May 2021, where it is suspected that a significant number of COVID-19 infections were not recorded.

#### 2.5.3 Model calibration sequence

To facilitate our work we have developed configurable software which uses Excel, MATLAB and Simulink. Simulink is used for the simulations which are optimised using MATLAB. The simulations and optimisations are managed from a configurable scripting system written in a structured Excel workbook which allows sequential strategies for model calibration and scenario forecasting to be defined.

MATLAB code interfaces with the Excel workbook. It reads and executes a series of optimisations defined by the instruction set. Each instruction set defines the data window for model calibration, the terms included in the objective function and the subset of model parameters selected for each optimisation stage. The initial conditions for the model parameters and initial estimates of optimisable parameters are also read from the Excel workbook. The initial estimates of optimisable parameters are used in the first optimisation stage only. For subsequent optimisation stages, the initial value of an optimised parameter is the final value from the previous stage.

After an initial trial and error procedure to define an appropriate initial temperature, the sequential approach used to calibrate the model to national UK data in both periods one and two is summarised in **Table 1**.

**Table 1.** Sequential method used to calibrate the model for UK data (crosses indicate active selections for the objective function and the subset of model parameters that are optimised at each stage). In stage 1, the first 60 days of data are used, specifying the objective function as the weighted sum of the cumulative dead, daily deaths and daily infections (weighting of 1 for daily infections, 1000 for daily deaths, and 100 for accumulated deceased individuals). In stage 2, the temperature co-ordinates across the entire data set were estimated keeping the estimated rate constants from the first optimisation stage constant. In stage 3, we re-optimise the kinetic model parameters using the first 95 days of data (data window chosen by trial and error to produce a good model calibration). The final stage then provides additional refinement of all the temperature co-ordinates to achieve the best possible model calibration. During model calibration all parameters are bounded as follows; 1*e*5 ≤ *A* ≤ 1*e*7, 0.001 ≤ (*γ*_*R*_, *γ*_*D*_) ≤ 0.2 and 273 ≤ *T* ≤ 373. The optimisation procedure uses jacketing of the optimisation algorithm to perturb the solution to allow optimisation to progress when local minima are encountered. As the value of *E* in equation **(13)** is a scaling factor for temperature in all model calibrations we fixed this value as, *E* = 5.859 × 10^3^ *K*, a value found in our previous modelling work (Willis, Wright, Bramfitt, & Diaz, 2021) when calibrating a model to reported case data.

| Stage | Time period<br>(days) |  | Objective Function |  |  | Optimised Parameters |  |  |  |
| --- | --- | --- | --- | --- | --- | --- | --- | --- | --- |
| | Start | End | Dead | Daily deaths | Daily infections | Rate constants | $A$ | $\gamma_R$ | $\gamma_D$ |
| 1 | Start | 60 | X | X | X | X | X | X | X |
| 2 | Start | End | X | X | X |  |  |  |  |
| 3 | Start | 95 | X | X | X | X | X | X | X |
| 4 | Start | End | X | X | X |  |  |  |  |

## 3.0 Results

The results of our model calibration for periods 1 and 2 are given in Table 2.

**Table 2:** Calibrated model constants for periods 1 and 2, UK national data and the London region

| | $A [d^{-1}]$ | $k [d^{-1}]$ | $\gamma_R [d^{-1}]$ | $\gamma_D [d^{-1}]$ |
| --- | --- | --- | --- | --- |
| UK (Period 1) | $1.78 \times 10^6$ | $9.02 \times 10^{-2}$ | $8.80 \times 10^{-2}$ | $2.22 \times 10^{-3}$ |
| UK (Period 2) | $1.78 \times 10^6$ | $1.70 \times 10^{-1}$ | $1.65 \times 10^{-1}$ | $4.82 \times 10^{-3}$ |

The values of *k* and *γ*_*D*_ show large differences between the original and Kent variants of COVID-19. The Kent variant has a value of *k* 88.9% higher than the original variant. As *k* is the specific transmission probability per exposure time, a constant that is characteristic of the COVID-19 infection, this implies that the Kent variant is more transmissible than the original variant. For the original strain, the ratio *γ*_*R*_: *γ*_*D*_ is 39.6, and for the Kent variant it is 34.2. Both these values are within the range of expected COVID-19 mortality rate of 1–4% (Joint Committe on Vaccination and Immunisation, 2021).

### 3.1 Calibrated model predictions of daily infections and deaths (Period 1 and 2)

Figure 1a and Figure 1b shows the actual and calibrated model predictions of daily infections and deaths using the entire UK data set. We performed model calibration separately for the data in periods one and two. Results from both calibration periods are combined in the plots. The results show a good agreement between actual and simulated data for both periods. The values of the coefficient of determination (*R*^2^) f or the number of infections were found to be 0.943 and 0.833 for the original and Kent variants. While for the deceased individuals they were found to be 0.999 and 0.998 respectively.

**Figure 1a.**
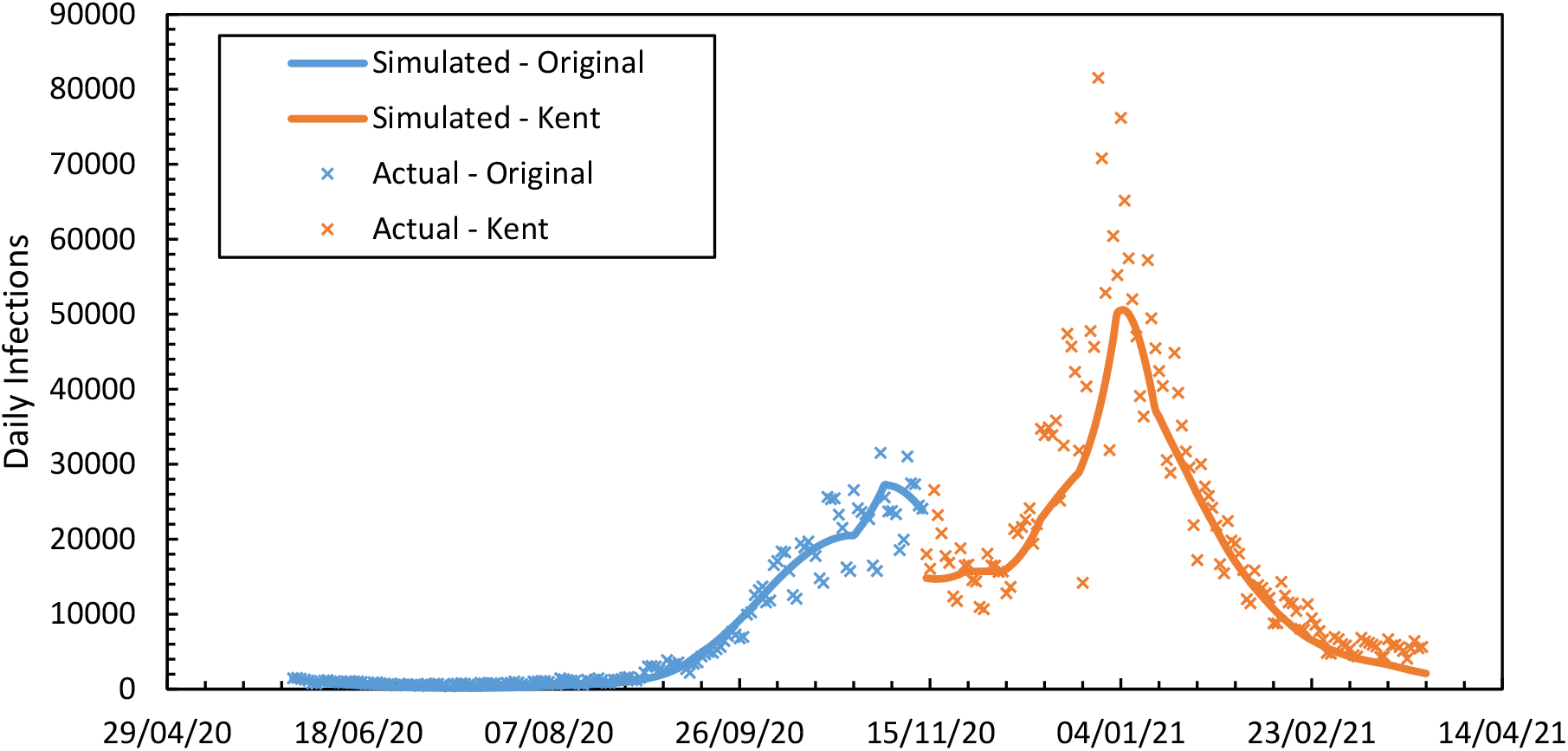
Daily COVID-19 infections in the UK combining results for periods one and two.

**Figure 1b.**
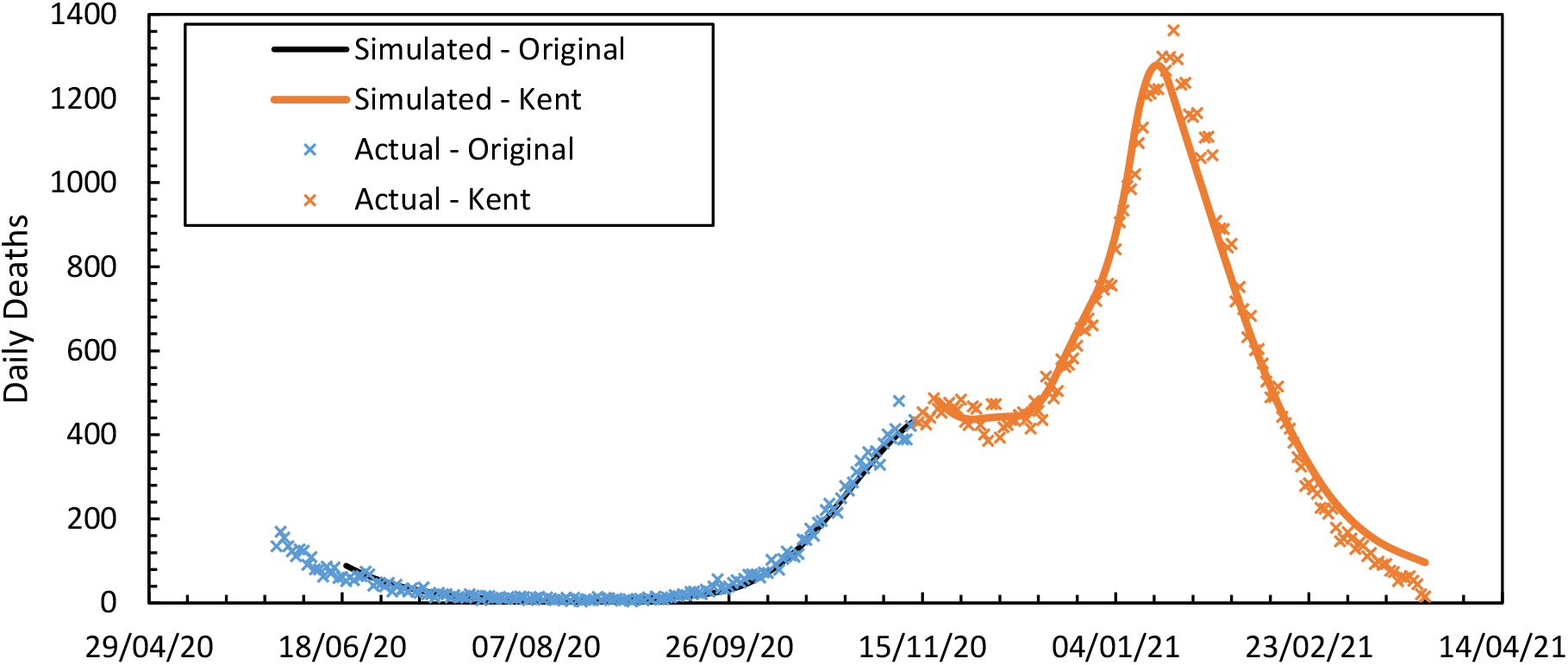
Daily COVID-19 deaths in the UK combining results for periods one and two.

**Figure 1a.**
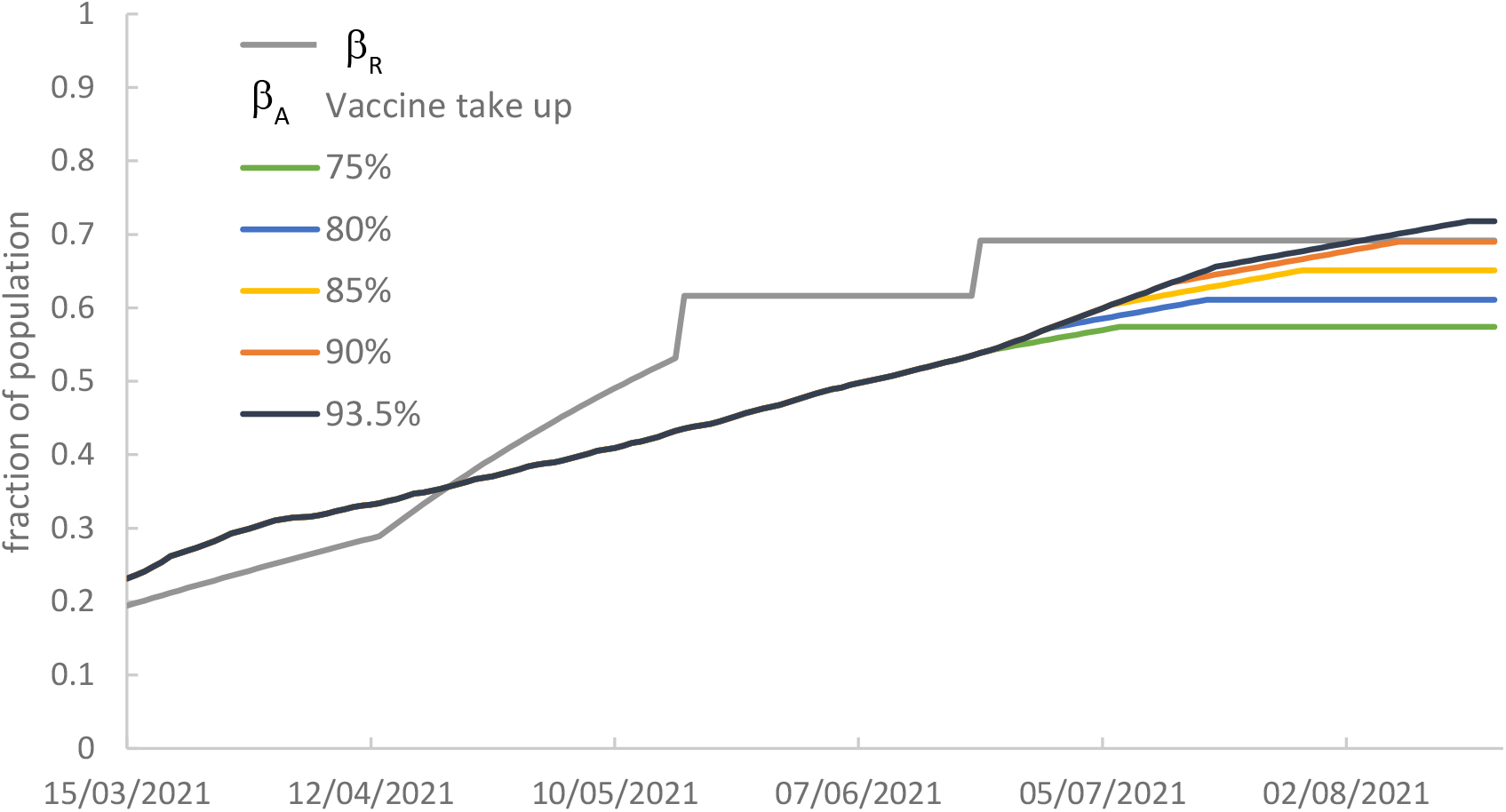
*-β*_*R*_ indicates the fraction of the population required to be immune from COVID-19 infection to ensure the number of infected individuals decreases. *β*_*A*_ is the actual fraction of the population immune from infection, estimated using the UK Government vaccine rollout (National Health Service, Statistics - COVID-19 Vaccinations, 2021c)

As shown by both the model and data, the number of COVID-19 infections had decreased to very low values by July 2020 following the first national lockdown. The number of infections remained low until September 2020, when the number of infections began to rise as the value of *R*_*e*_ increased above one. The number of infections across the country remained stable in November 2020 because of the second national lockdown. This lockdown was shorter and implemented fewer NPI restrictions than the first and third lockdowns (BBC News, 2021), which prevented the number of infections decreasing further. Following the second national lockdown a local tier system was reintroduced. This introduced more restrictions than the previous tier system. However, the measures were not sufficiently stringent to prevent increased transmission and the number of daily infections reached a peak in January 2021. The UK entered its third national lockdown at the beginning of 2021, and the strict restrictions placed on the UK population during this lockdown resulted in the number of infections decreasing sharply.

Generally, the trend of daily COVID-19 deaths reflects the number of daily infections with a 2-week lag. There are notable and clear similarities between the two figures, however there are some exceptions to the general trends. For example, the number of infections remains stable in November 2020, whereas the number of daily deaths decreased during the same time. One possible explanation is that the increased number of COVID-19 tests being performed resulted in more infections being detected and reported.

The values of *R*_*e*_ in the profile produced by the model for both the original and B.1.1.7 (Kent) variants of COVID-19 are also shown in Figure 1c. The UK Government published an upper and lower estimate of the value of *R*_*e*_ in the UK throughout the pandemic, which is also shown in the Figure 1c. There is a clear correlation between the results of our model and the official data. It may be observed that the *R*_*e*_ profile of the original variant of COVID-19 produced by our model has larger minimum and maximum values than the official data. The maximum value of *R*_*e*_ produced by our model is 2.0, compared to 1.6 in the official data. When our model was calibrated to the B.1.1.7 (Kent) variant of COVID-19 it produced an *R*_*e*_ profile which remained closer to the range estimated by the UK Government.

**Figure 1c.**
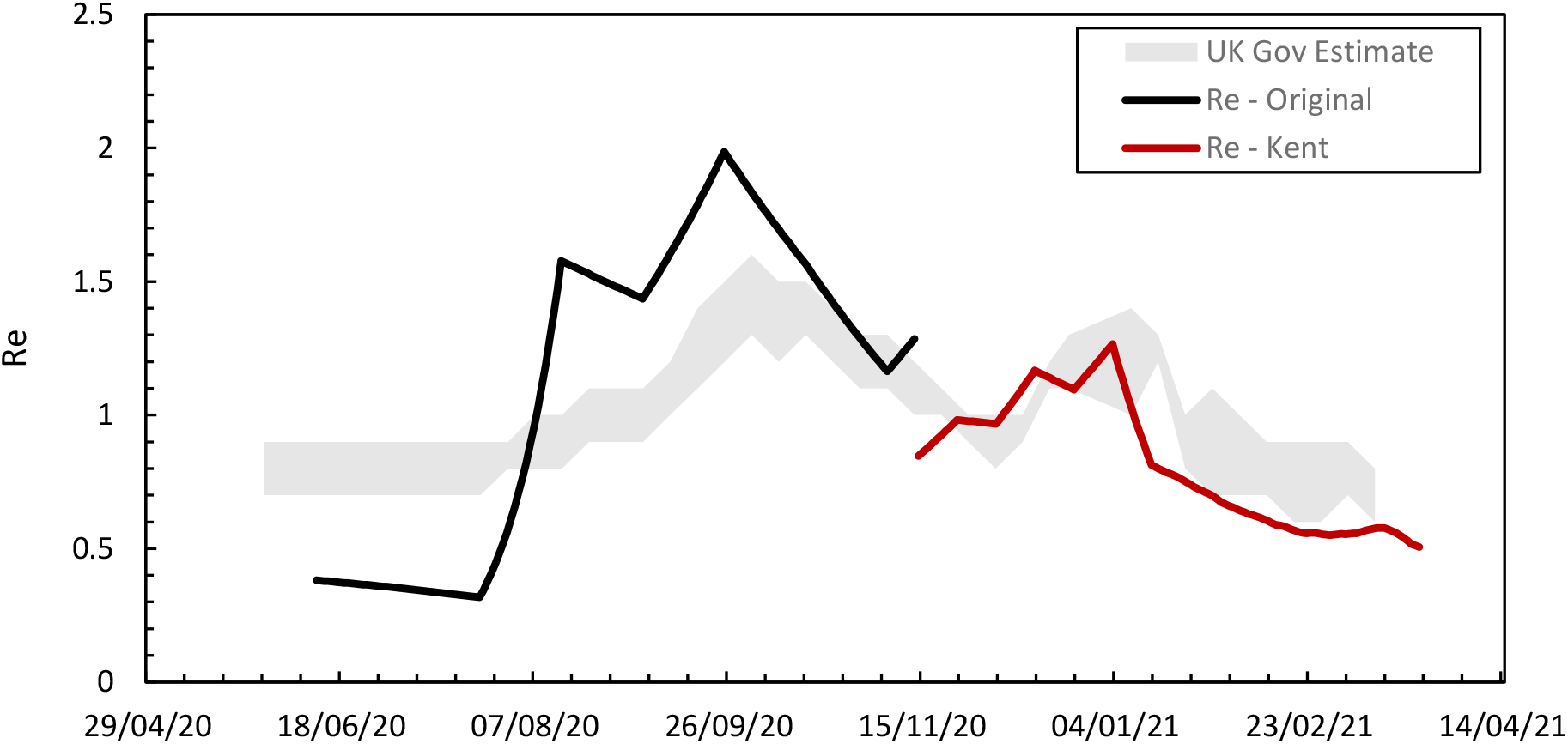
The calculated value of R_e_ over time combining results for periods one and two compared to the estimated Government range.

### 3.2 Model calibration: UK data between March 2020 and May 2021

As community testing for COVID-19 was not implemented by the UK Government until May 2020, it is estimated that the vast majority of COVID-19 infections were unreported before this date. However, as COVID-19 tests were carried out on people seriously ill in hospital, it can be assumed that the majority of COVID-19 deaths were reported. The original variant of COVID-19 was present in the UK between March 2020 and May 2020 (Chowdhury, Scarr, MacAskill, & Marshall, 2021), and therefore the kinetic model parameters calculated for the UK original variant (Table 2) are assumed to be applicable to the spread of COVID-19 during this time. We calibrated the model for UK data between March 2020 and May 2020 using only the number of daily and cumulative deaths as objective functions in order to estimate the number of infections within this time period. To perform model calibration, the set of temperature co-ordinates which were defined every 10 days were adjusted.

The results are shown in **Figure 2a - c** (*R*^2^ = 0.981 for the daily death estimation). The number of daily COVID-19 infections found by the model was significantly higher than the data published by the UK Government. Using the difference between the simulated and published daily infections, it is estimated that ∼1.2*M* COVID-19 infections were unreported between March 2020 and June 2020. Additionally, we find the basic reproduction number to be *R*_0_ = 3.23, a value comparable to many initial estimates which were in the range *R*_0_∼3.0 − 3.9 (Locatelli, Trächsel, & Rousson, 2021) (Lui, Tang, & Lam, 2020) (Bruce, et al., 2020). We use this estimate of the basic reproduction number to assess the final stages of the UK government road map due to occur on the 21 June 2021.

**Figure 2a.**
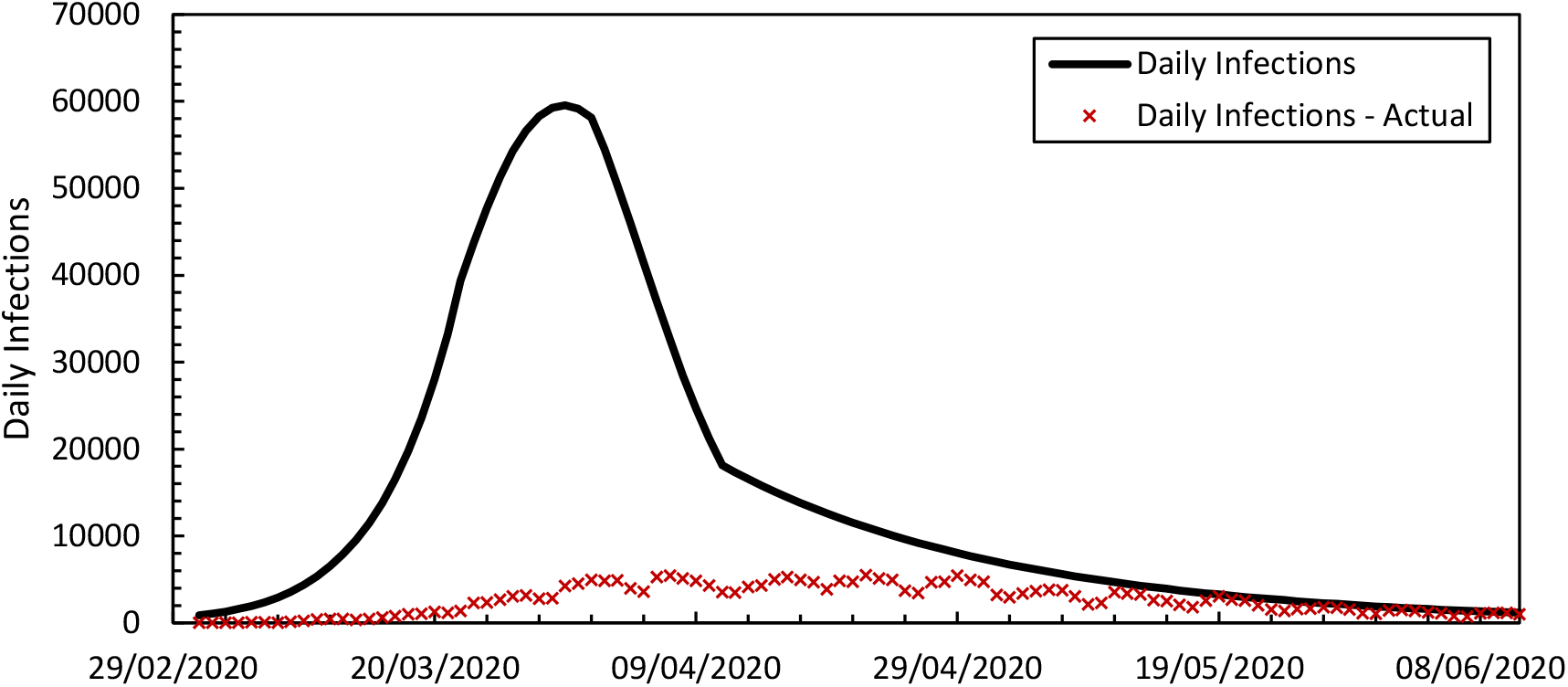
Daily COVID-19 infections in the UK. The solid lines show the simulation results, and the crosses show the reported case data. The calibrated model shows the significant under reporting of cases in the early stages of the UK epidemic.

**Figure 2b.**
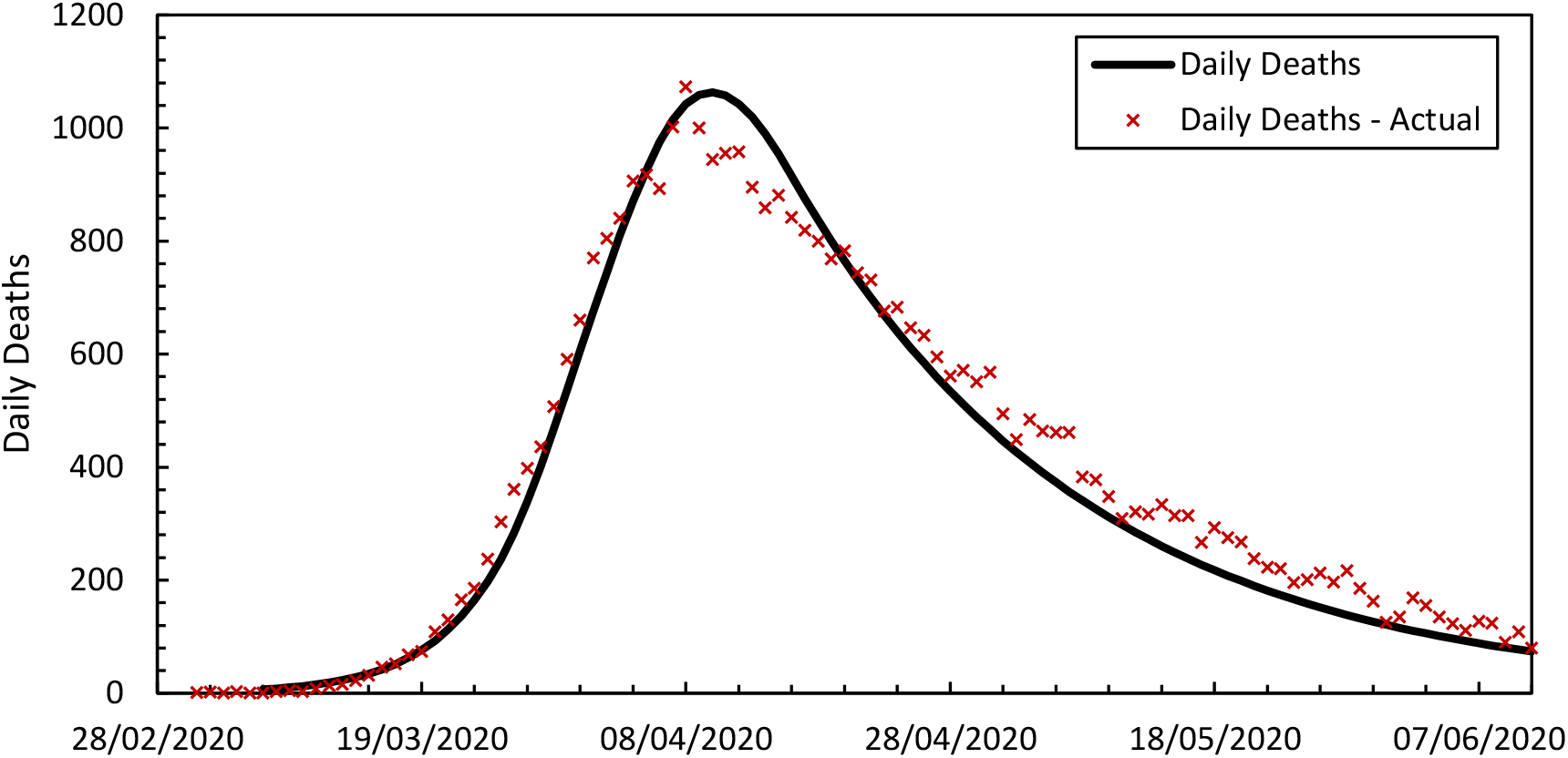
Daily COVID-19 deaths in the UK

**Figure 2c.**
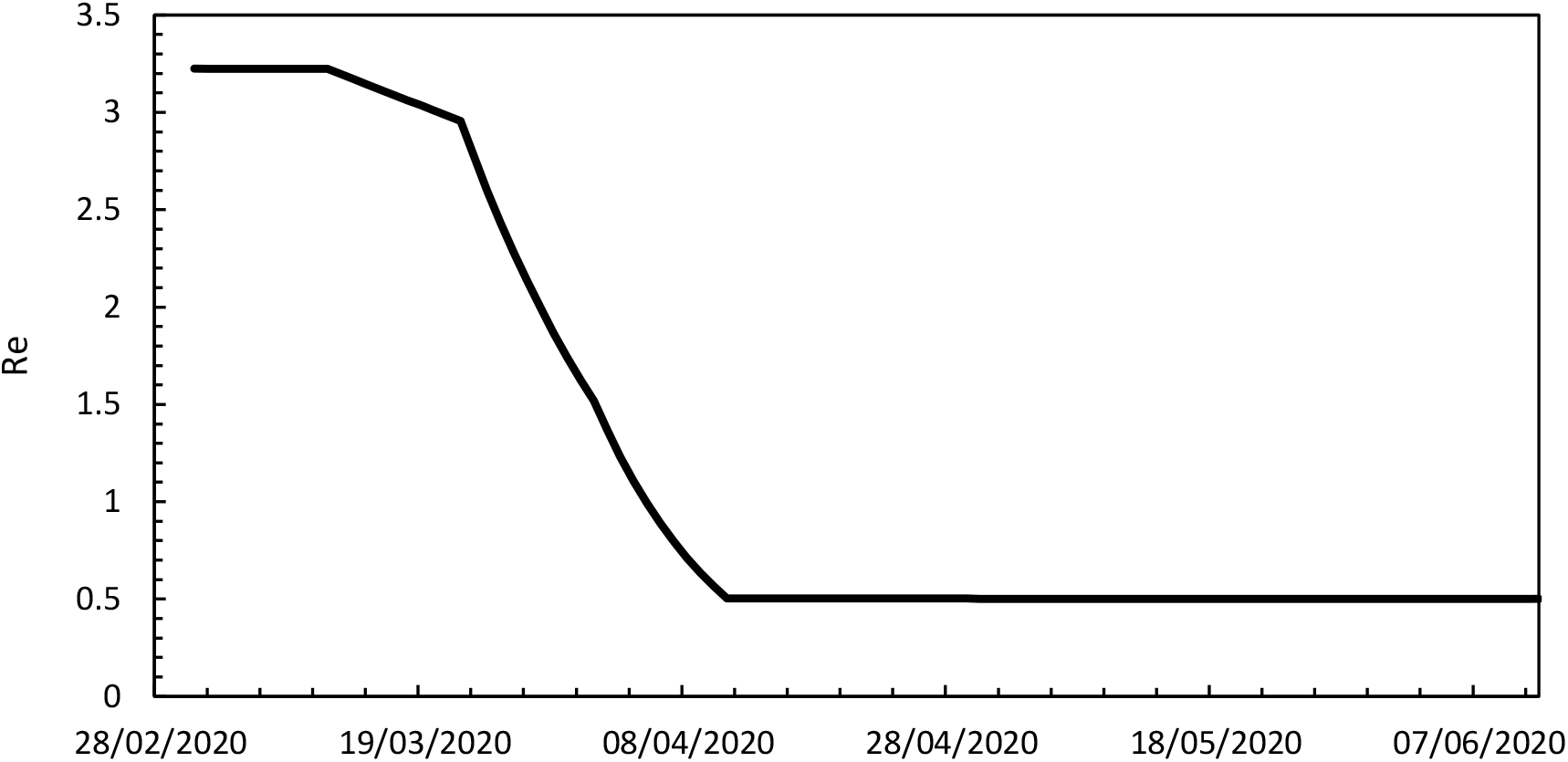
Value of *R*_*e*_ over time from March to June 2020.

**Figure 2b.**
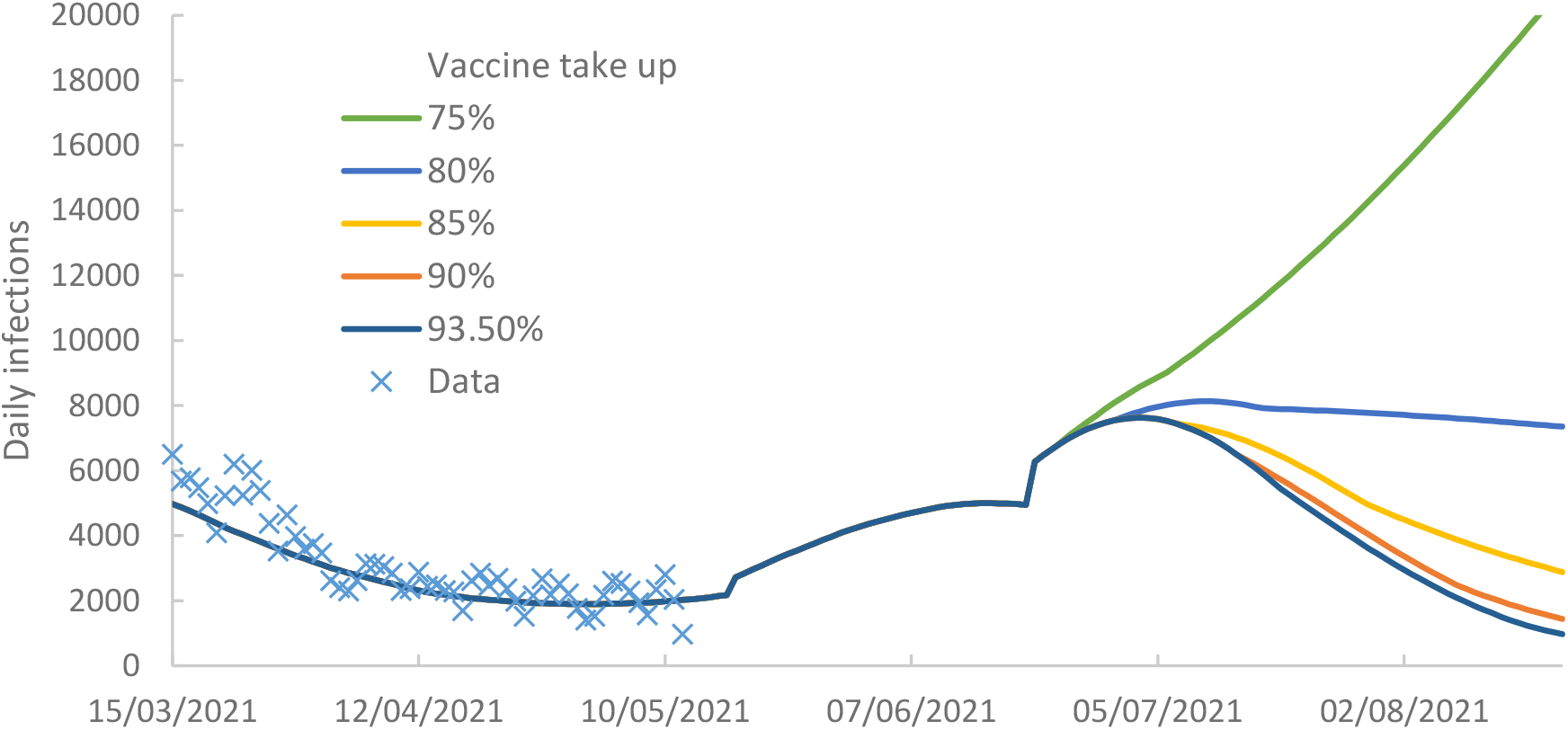
Forecast daily infections for different levels of vaccine take up

### 3.3 Vaccine roll-out, herd immunity and the requirement for future NPIs

We use equation **(23)** to assess the effectiveness of the vaccine roll-out and potential future scenarios as NPIs continue to be relaxed according to the UK Governments road map, where further lifting of restrictions are currently scheduled to take place on the 17 May and 21 June 2021. Until 16 May 2021 we use the value of the effective reproduction number calculated by the simulator. *R*_*e*_ increases gradually, reaching 2.3 by 16 May 2021. A step increase to 2.6 is imposed on 17 May 2021 to correspond with the release in lockdown measures. This is the upper limit of the range of values suggested by the Imperial College COVID-19 Response Team. (Imperial College COVID-19 Response Team, 2021). For the scheduled easing of NPIs in June 2021 we use our estimated value of *R*_0_ = 3.23.

The vaccination forecast assumes a total daily vaccination capacity of 500,000 which is slightly greater than the average number of daily vaccinations achieved in the preceding two months and in line with the Government’s objective of increasing vaccination rates. On a given day, that the number of second vaccinations corresponds to the number of first vaccinations given 12 weeks previously and assumes that all second vaccinations offered are taken up. The number of first vaccine doses given each day is calculated as the difference between the number of second doses given and the daily capacity. First vaccinations are given until the number of eligible individuals willing to be vaccinated is reached. For example, the maximum number of individuals receiving a COVID-19 vaccine will be (78.6% eligible) · (93.5% take-up) · *N* = 49,890,108. Vaccine efficacy is assumed to be 67% for the first dose and 98% for the second dose. It is assumed that level of immunity for vaccinated individuals reaches the maximum efficacy of the dose immediately and does not decrease over time.

In Figure 3a, the value of *β*_*A*_ is less than the value of *β*_*R*_, between 21 April 2021 and 4 August 2021. There is potential in this period for the number of infections to rise. *β*_*R*_ rises sharply as the third national lockdown is eased according to the UK Government’s roadmap.

Although *β*_*R*_ increases, the fraction of the population immune from COVID-19 due to the rollout of vaccines continues to increase. At 93.5% take up of vaccination, *β*_*A*_ increases so that it is greater than *β*_*R*_ in August 2021 and herd immunity is achieved.

At lower levels of vaccine take-up, *β*_*A*_ remains less than *β*_*R*_ prolonging the period where a surge in infections is possible. The simulation results shown in Figure 3b quantify the increase in daily infections for a given vaccination take up. At the present level of 93.5% take up, there is a small increase in daily infection numbers in May. At the time of writing (mid May 2021) the numbers of daily cases are showing a slight upward trend.

A more significant increase in new infections is forecast in June 2021 with a peak in excess of 7000 new daily cases. This increase corresponds to the planned lifting of all NPIs in the Government roadmap. If the vaccination take-up rate falls as younger age groups become eligible for vaccination, the forecast surge in new daily cases rises, and a vaccine take up of less than 80% may result in loss of control of number of new cases.

## 4.0 Discussion and conclusions

We have demonstrated the effectiveness of using parameter regression methods to calibrate an augmented *SIRD* model that includes vaccination dynamics. Our results indicate that our model, where *R*_*e*_ varies exponentially as a function of NPIs, can accurately capture the reported daily case numbers. We have used this model to explore the characteristic dynamics of the COVID-19 epidemic in the UK. Understanding that in any *SIRD* model *R*_*e*_ is a variable stochiometric coefficient in the infection step has enabled the determination of *R*_*e*_ · *k* by model calibration. This approach also allows numerical decoupling of *R*_*e*_ and *k* which has allowed us to detect a change in the characteristic rate of infection as a result of the COVID-19 mutation colloquially known as the Kent variant. Moreover, through retrospective analysis of reported case data we have been able to determine an estimate of the basic reproduction number as, *R*_0_ = 3.23.

We have used our model to assess the UK Government’s roadmap for easing of the third national lock-down. To do this we used our calibrated model parameters, estimates of the effective reproduction number at key stages of the easing of the lockdown and a vaccination rate forecast for the coming months. Our work does not consider data beyond 12 May 2021 and therefore is confined to the B.1.1.7 (Kent) variant only. Contemporaneous reports of a virus variant of concern (the Indian variant B.1.617.2) are emerging. This strain is possibly more infectious however it is not considered in our analysis. We also note that immunity is not fully developed for several weeks after vaccination, which will extend the period where there is potential for a surge in infections.

- Our forecast of infection numbers assume *R*_0_ = 3.23 from 21^st^ June 2021. It is unlikely this value will be achieved as other measures will remain in place for example wearing of masks and social distancing etiquette is still likely to occur.
- The model does indicate that there may be an increase in case numbers. This is anticipated by the UK Government (H M Government, 2021) which proposes increased testing in educational establishments and workplaces, surge testing, contact tracing and isolation of infected individuals.
- The plot of *β*_*A*_ and *β*_*R*_ in Figure 3a provides an indication of the period where the numbers of vaccinated individuals are insufficient to supress growth in case number. This analysis is subject to our assumptions for *R*_*e*_ on lifting of NPIs and for the vaccination programme. It is clear that when *β*_*A*_ < *β*_*R*_ the number of cases may increase. There is some uncertainty in the forecast however there is a clear nonlinear relationship between the daily rate of new infections shown in Figure 3b and the magnitude of *β*_*R*_ − *β*_*A*_ in Figure 3a.
- It is apparent that reduced vaccination take up in eligible adults may lead to a rapid increase in case numbers.

## Data Availability

All data used is available in the public domain from the sources referenced in the manuscript.

## Footnotes

1 deterministic, autocatalytic kinetic models of the whole population (Alexandru & Ross, 2015)

2 This is available within MATLAB’s optimisation toolbox

3 As many services and offices close on weekends in the UK, the number of new COVID-19 infections and deaths registered can drop by up to 50% on weekends when compared to weekdays (Ricon-Becker, Tarrasch, Blinder, & Ben-Eliyahu, 2020).

